# Impacts of Early Interventions on the Age-Specific Incidence of COVID-19 in New York, Los Angeles, Daegu and Nairobi

**DOI:** 10.1101/2020.04.19.20071803

**Authors:** Hao Yin, Zhu Liu, Daniel M. Kammen

## Abstract

**Background:** COVID-19 has caused an unprecedented public health crisis and economic shock to the global economy. While many countries were affected, regions with an older population and weaker public health interventions tended to suffer more morbidity and mortality. Here we model and quantify the age-specific incidence of COVID-19 in four pandemic cities under different interventions.

**Methods:** We developed an age-specific and multiple-stage susceptible-exposed-infected-recovered-hospitalized-quarantined-dead (SEIR-HQD) dynamical systems model expanded from the more basic SEIR model by incorporating location- and age-specific contact matrices to estimate the outcomes of COVID-19. Utilizing latest estimates of epidemiological parameters and demographic data, we model the potential effects of various interventions in four representative cities with different population structures - New York, Los Angeles, Daegu and Nairobi. We compared the effects of different interventions in the age-structure populations specific to each city. These policy options are then applied to determine the potential for effective containment. We model these dynamic policy scenarios to assess the risks of less-stringent social distancing, as has been proposed by those arguing to enhance economic activity over public health and safety. Finally, we explored the health impacts of different policy action timelines to understand the benefits of early interventions.

**Findings:** We find the spread of COVID-19 to be dramatically different in the regions modeled, with the primary drivers the variation of population age structures, and the dynamics of interactions of the younger demographics, whose higher interaction rates can lead to increasing transmission rates across these communities. A city with younger citizens may also have fewer hospitalized cases and deaths. Our modeling quantifies the value of early interventions, which avoided an additional 5%, 16%, 37% and 43% of the infections in Daegu, Nairobi, New York and Los Angeles, respectively, compared to what has been observed in the four cities. The finding is clear: in the absence of pharmaceutical options, delaying strict social policy interventions has resulted in substantial public health cost. This modeling can, and will, be applied to other cities and regions, and conducted in conjunction with other health insults, such as exposure to air pollution.

Critically, we find that school closures, working from home, and reduction in other mobility were most beneficial for younger population (0-19 years old), middle-age (20-59 years old) population and older population (60 years and older), respectively across each city. Specifically, school closure avoided 25%, 18%, 16% and 12% of the infections for the population under 20 years old in Daegu, Los Angeles, New York and Nairobi, respectively. A 50% and 80% population working from home policy avoids 8% and 15% of the infections. Reduction in mobility was more effective than the working from home strategy. Any single social distancing policy if enacted alone can delay the spread of COVID-19 but was unable to totally suppress the infection. Coordinated policy action can be highly effective. Increasing the quarantine rate to 10% of infectious cases was more effective than strict social distancing alone in this study, although together they can suppress 80% of the epidemic. A combination of moderate social distancing and quarantine strategies was able to avoid 99% of the infections.

**Interpretation:** Moderate social distancing together with high quarantine rates was effective in each of the four cities. COVID-19 caused more deaths and hospitalization in cities with an ageing population than those with a younger population. However, in the cities with a younger population, there is a clear need to implement a social distancing strategy that is even more strict due to the higher transmission rates among younger people. Cities with more older people should prepare more hospital beds and healthcare facilities to save people who are in critical conditions. Cities with ageing population should take targeted action for the elderly to avoid the severe impacts on the vulnerable populations. Increasing quarantine rate is an effective strategy to avoid the substantial infection while also does not influence the economy fiercely. We recommend countries or regions experiencing, or likely to experience the rapid spread of COVID-19, to implement combination of multiple strategies in the early stage of the breakout which can avoid over 90% of infected cases.

**Funding:** National Natural Science Foundation of China, China Postdoctoral Science Foundation, Qiushi Foundation and the Resnick Sustainability Institute at California Institute of Technology, Zaffaroni Family Foundation, the Karsten Family Foundation, the National Science Foundation of the United States.

## Introduction

The outbreak of COVID-19 triggered heterogenous responses from countries across the world. While the effectiveness of these containment strategies is still emerging, and critical lessons for how long social isolation and testing must remain in place, as well as the need to guard against re-infection demand immediate access to models that can be ground-truthed by comparing model results and the emerging responses to policies already in place in diverse urban settings. For the most hard-hit countries, such as the US, Italy and Spain, there are greater percentages of older population who are facing much higher risks of hospitalization and fatality due to changes of immune system and the impact of preexisting health conditions over time.^1^ Studies have found that COVID-19 was an emergent disease of ageing.^2^

The effects of population ageing on the outcomes of COVID-19 require much more resources in healthcare services and facilities, which is places a significant burden on the health care infrastructure of many ageing countries. Therefore, countries with high proportion of older population should prioritize strategies that effectively protect older people during the epidemic. The difference in demographic characteristics may call for targeted interventions to suppress the spread of COVID-19. Studies have emphasized the importance of incorporating the demographic information in curbing the spread of the epidemic.^3^ An important recent study by the Imperial College COVID-19 Response Team explored the impacts of public health interventions in 11 European countries.^4^ Here we focus on individual cities where the policy steps are more homogeneous and can be well-described in the model we present and make available here. Understanding the effects of population ageing on the spread of the epidemic under various containment interventions enables policymakers to make effective decision making to avoid massive hit of the novel coronavirus and relief the pressure on healthcare systems.

Important and critically needed modeling studies have explored the effects of different interventions, such as social distancing, school closing and managing dynamics of population movement.^5–9^ Social distancing has been recognized as one of the most effective mitigation strategies.^10,11^ An under-studied issue is that of mental health impacts of these strategies across different segments of the population. In addition to social distancing, there are additional opportunities for countries to make transition from stopping the economy. Strategies to get young people back to work and protect older population and epidemiological surveys to track the exposed cases and strict quarantine are implemented well in Korea and Singapore. Strategies tailored to different age groups within the population are in an urgent need to recover the economy.

In this study, we focused on four cities, New York, Los Angeles, Daegu and Nairobi, that both shared similarities and reflected diversities in terms of demographics and containment strategies. As the world’s most impacted nation, the number of COVID-19 cases in the United States (US) surged to over half a million by mid-April.^12^ New York, the most populous US metropolitan area, was hit the hardest. By contrast, Los Angeles, had a total number of infected cases accounting for fewer than 10% of that in New York. Many factors influence such differences, but the timeline of policy interventions in Los Angeles and New York indicates that New York took actions even faster since the detection of the first case. However, the intervention policy is less detailed and instructive for the public to follow than the one in Los Angeles. Explanations for the rapid increase in the spread of COVID-19 have been proposed.^13,14^ Compared with the containment strategy in the US overall, Korea implemented a faster and effective testing regime, contact-tracing, and quarantining of people. Detailed online education and clarification of requirements for isolation where widely disseminated online. In total the South Korean policies greatly increased the quarantine rate of exposed population. Nairobi, as the capital city of Kenya, has a relatively younger population than the other three cities. Kenya also take a fast response since the first case was detected. Four cities all ordered people to keep social distancing, close schools and hand hygiene. While Korea and Kenya called people to wear face masks in the beginning, the US did not encourage people to wear face masks/cloth face covering until April 3, 2020.^15^ The difference in the interaction of these non-pharmaceutical interventions and age structures of population might deviate the spread of COVID-19.

## Research in context

### Evidence before this study

We searched PubMed, google scholar, medRxiv for studies on the epidemic modeling of COVID-19 published up to April 17, 2020, using the search terms “COVID-19”, “2019-nCoV”, “SARS-CoV-2” with “epidemic modeling”, “older population”. Many studies have modelled the epidemic trends and healthcare impacts of COVID-19 using SEIR or other mathematical models. These models are applied to predict the epidemic peaks and sizes of COVID-19 in different intervention scenarios, such as closing schools, working from home, household quarantine and city lockdown. Age-specific studies have reported the role of school children on the spread of COVId-19. Some studies also highlighted the vulnerability of older population with pre-existing conditions. These studies mainly focused on the overall population, while it remains unknown about the effects of population demographics on the spread of COVID-19.

### Added value of this study

We developed an age-specific SEIR-HQD model based on the contact matrices across population in different age groups. In this study, we explicitly explored the age-specific effects of different non-pharmaceutical interventions on the spread of COVID-19. We found that strategies related to social distancing had strong age-specific effects on the epidemic peak and sizes. Therefore, cities with more ageing population should have targeted strategies to protect them. While older population was vulnerable, the younger population was more transmissible than the older individuals. A younger city might have more infected cases whereas fewer deaths and hospitalized cases than an ageing city. We found that 1-week earlier interventions were more effective in Los Angeles and New York which indicated that these two cities might take actions slower than Daegu and Nairobi according to recent policymaking on COVID-19. Sustained policy intervention was helpful to avoid a second wave in the four cities, especially in Nairobi.

### Implications of all the available evidence

General social distancing intervention could help to contain the outbreak of COVID-19. More specific social distancing strategies aiming at protecting the elderly are essential for cities with an ageing population. Moderate social distancing among the younger population and a high quarantine rate could help to contain the rapid spread of COVID-19. Cities with younger population should implement a stricter social distancing intervention than an ageing city. Whereas, cities with older population should prepare more healthcare facilities and hospital beds.

## Methods

### 1. Epidemiological modelling of COVID-19 outbreak in four cities

In this study, we expanded a SEIR model to an age-specific SEIR-HQD model for a period of 365 days. SEIR-HQD model considers 7 states of infection including susceptible (S), exposed (E), infectious (I) and recovered (R), hospitalized (H), quarantined (Q) and dead (D). The SEIR-HQD model incorporates the age structure, birth rates and death rates dynamics of the affected population in Daegu, New York, Los Angeles and Nairobi.

We stratified each population into 9 age groups by a 10-year band, with the last age group set as those 80 years and older. We assume the population in these four cities are closed systems without population flow inside or outside the cities. When the susceptible populations are exposed to infectious individuals, a percentage then transition into infected status at a given probability defined by an age-specific transmission rate. After the incubation period, the exposed population then become infectious, but if they are quarantined, we assume that they do not impact susceptible individuals in the next generation. For those who are infectious, given the hospitalization rate of infectious cases, we further calculate the number of people who are in need of further medical care in the hospital. Our model also simulates the quarantine of individuals and hospitalized cases are sub-states of infected cases that will not cause a secondary infection. The quarantine individuals might also become hospitalized. The epidemic evolution model is described as follows:

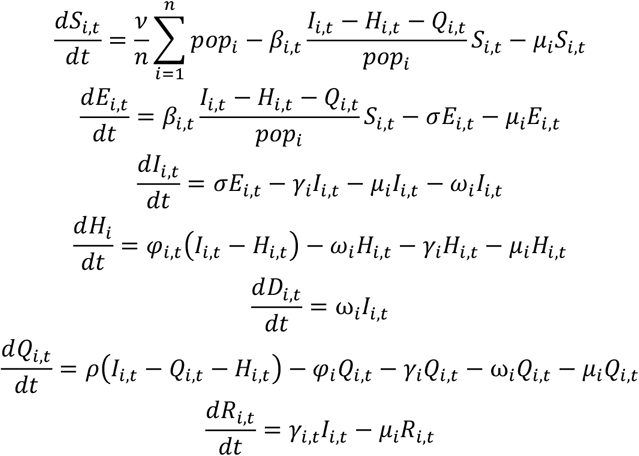

*S*_*i,t*_ is the susceptible population in age group *i* on day *t*,

*v* s the birth rate per thousand population,

*μ*_*i*_ is the death rate of population in age group *i*,

*n* is the number of age groups, *pop*_*i*_, is the number of population in age group *i*,

*β*_*i,t*_ is transmission rate, defined as the probability of infection between a susceptible and infected individual,

*I*_*i,t*_ is the infected people in age group *i*,

*E*_*i,t*_ is the exposed population in age group *i*,

*σ* is the daily probability of an exposed individual becoming infectious, which equals to 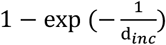, where d_*inc*_ refers to duration of average incubation time,

*ρ* is quarantine proportion of exposed individuals,

*γ*_*i,t*_ is the probability of an infected individual that recovers during the infectious duration d_*inf*_, so that 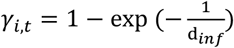,

*φ*_*i,t*_ is the hospitalization rate in age group *i*,

*Ω*_*i,t*_ is the fatality rate of infected individuals,

*Q*_*i,t*_ is the number of individuals that are quarantined,

*R*_*i,t*_ is the number of infected individuals who recover.

The parameters involved in the model are obtained from literature and are presented in Table 1.

**Table 1.** Parameters in the age-specific SEIR-HQD model

| Parameters | Value | Reference |
| --- | --- | --- |
| Basic reproduction number $R_0$ | 2.92 (2.28, 3.67) | Liu et al. (2020)<sup>16</sup> |
| Transmission rate | Varies by age, see Appendix 1. | Estimated in this study |
| Average incubation period, $d_{inc}$ | 6.4 days | Backer et al. (2020)<sup>17</sup> |
| Average duration of infection, $d_{inf}$ | 7-11 days (assume growing by age) | Cao et al. (2020)<sup>18</sup> and Bi et al. (2020)<sup>19</sup> |
| Initial number of infected, $I_0$ | 200 or 500 per million | Abbott et al. (2020)<sup>20</sup> and Prem et al. (2020)<sup>9</sup> |
| Hospitalization rate, $\varphi_{i,t}$ | Varies by age, see Appendix 1 | Ferguson et al. (2020)<sup>7</sup> and Verity et al. (2020)<sup>1</sup> |
| Quarantine rate, $\theta$ | 0%-80% | Assumed in scenarios |
| Fatality rate, $\omega_{i,t}$ | Varies by age, see Appendix 1. | Verity et al. (2020)<sup>1</sup> |

### 2. Transmission rate matrix and population age structure

The transmission rate differs from the location, population mobility and contact probability. Therefore, we calculate the transmission rate matrix based on contact matrices in different location settings at country level. We first adjusted the contact matrices by age-specific population, the more population in certain age group, the higher probability of this group population gets in contact with one in other age groups. Then we estimate the transmissibility using method:^21^

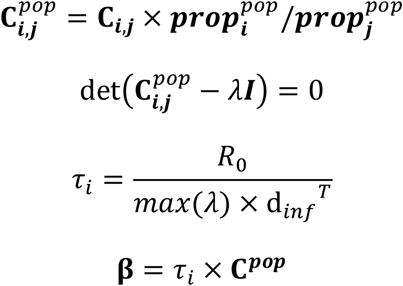

Where *i* and *j* represent age groups, **C**^*pop*^ is contact matrix in each city, 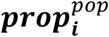 is the proportion of population in age group *i* to the total population in a city, *τ*_*i*_ is the transmission probability of a contact between an infectious individual with a susceptible one, *R*_0_ is the basic reproduction rate, and **β** is the transmission rate matrix among individuals in different age groups.

In addition to factors such as population density and social habits, the population age structure also influences transmission rates among different age group population. Nairobi has a younger population than the other three cities, thus there are lower transmission rates among older people and higher transmission rate among the younger people than that in the relatively ageing cities in the US and South Korea (Figure 1). The population age structure information is listed in the Appendix 1.

**Figure 1.**
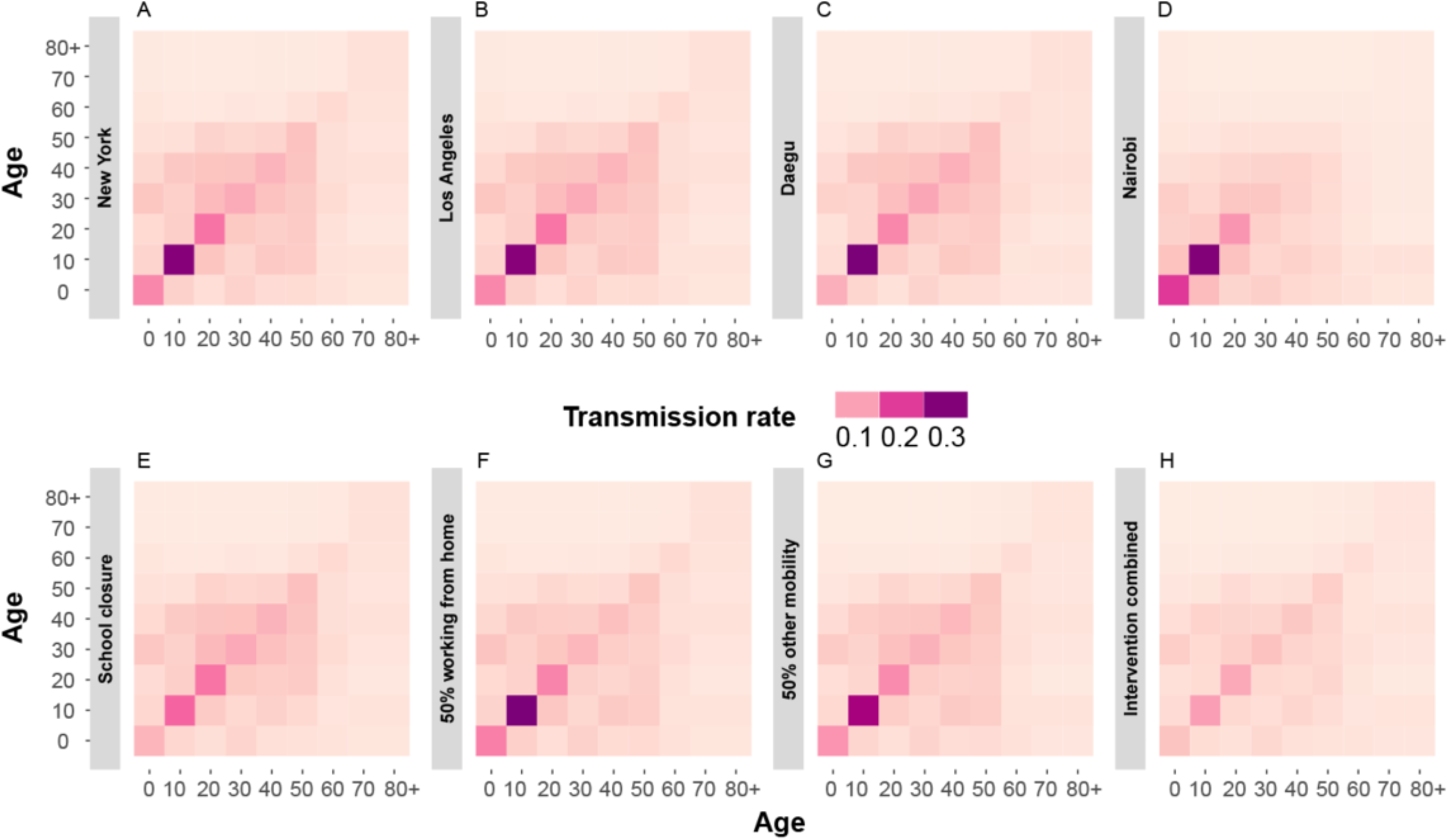
Transmission rate matrices in different cities (Figure 1A-1D) and different counterfactual transmission rate matrices in different intervention scenarios (Figure 1E-1H)

The timeline of the policy interventions illustrated the speed of reaction to suppress the epidemic peaks in each city (Figure 6). Since Nairobi was hit later than other three cities, we can forecast the effects of implemented policy interventions, and outcomes, may evolve significantly from what we have modeled here.

### 3. Scenario design for interventions and the timing of policy action

Applying the SEIR-HQD model, we designed two sets of scenarios to explore: 1) the age-specific effects of policy interventions and 2) the effectiveness of policy action at different time points since the diagnosis of the first case.

For the age-specific effects, we designed 7 scenarios that tested school distancing, reduction in other mobility, increasing quarantine rates and a mixture policy package.

We also reviewed the policy actions in four cities from the first case detected to April 9, 2020 (Figure 2). We followed the real policy actions to simulate the business-as-usual scenarios in the four cities. Then we move each strategy 1 week earlier to see the effects of early actions.

**Figure 2.**
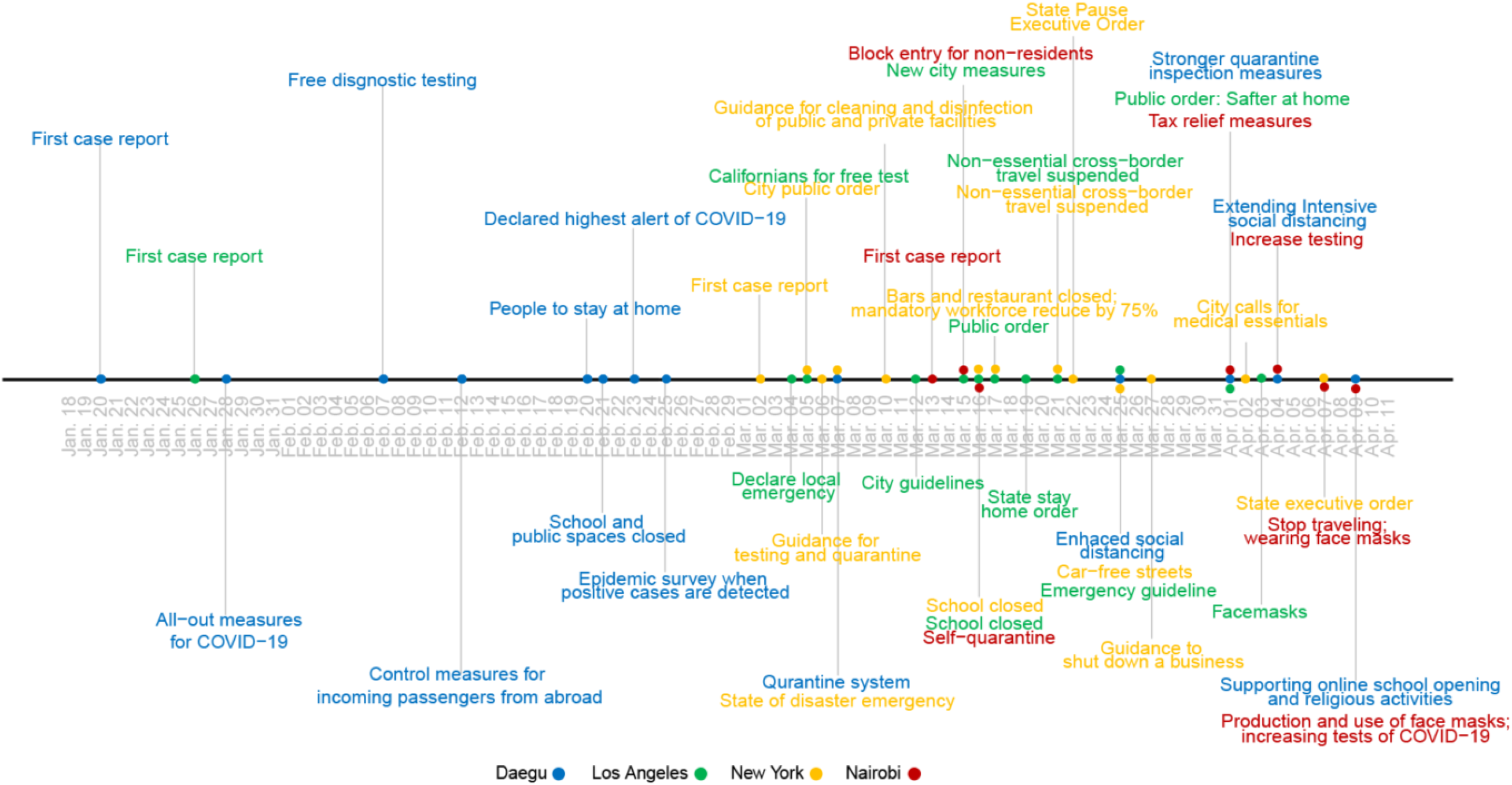
Timeline of policy interventions and first case reports in four cities, Daegu, South Korea, Los Angeles, USA, New York, USA and Nairobi, Kenya. Detailed information on the interventions summarized here is provided in Appendix 1.

## Results

### 1. The age-specific effects of different interventions on the outcomes of COVID-19

Figure 3 presents the temporal evolution of the number of individuals in each wellness/illness category (“status”) over a year period in the four cities simulated per million residents of initial susceptible population with no intervention. It shows that the COVID-19 outbreak hits the four cities dramatically although differently, mainly due to the variation of population age structures. In the ‘no intervention’ scenario, COVID-19 causes 550,000 infected cases per million population in Nairobi, which is 35% higher than the reported infected cases in New York, Los Angeles, Daegu. This was mainly due to the higher contact rates among younger population was relative to the older population, which caused higher transmission rates among them. However, the deaths related to COVID-19 were much lower than other three cities with older populations. As seen in Figure 3, for each million of the susceptible population, COVID-19 caused 29% more deaths in Daegu than in Nairobi, largely due to the older demographics.

**Figure 3.**
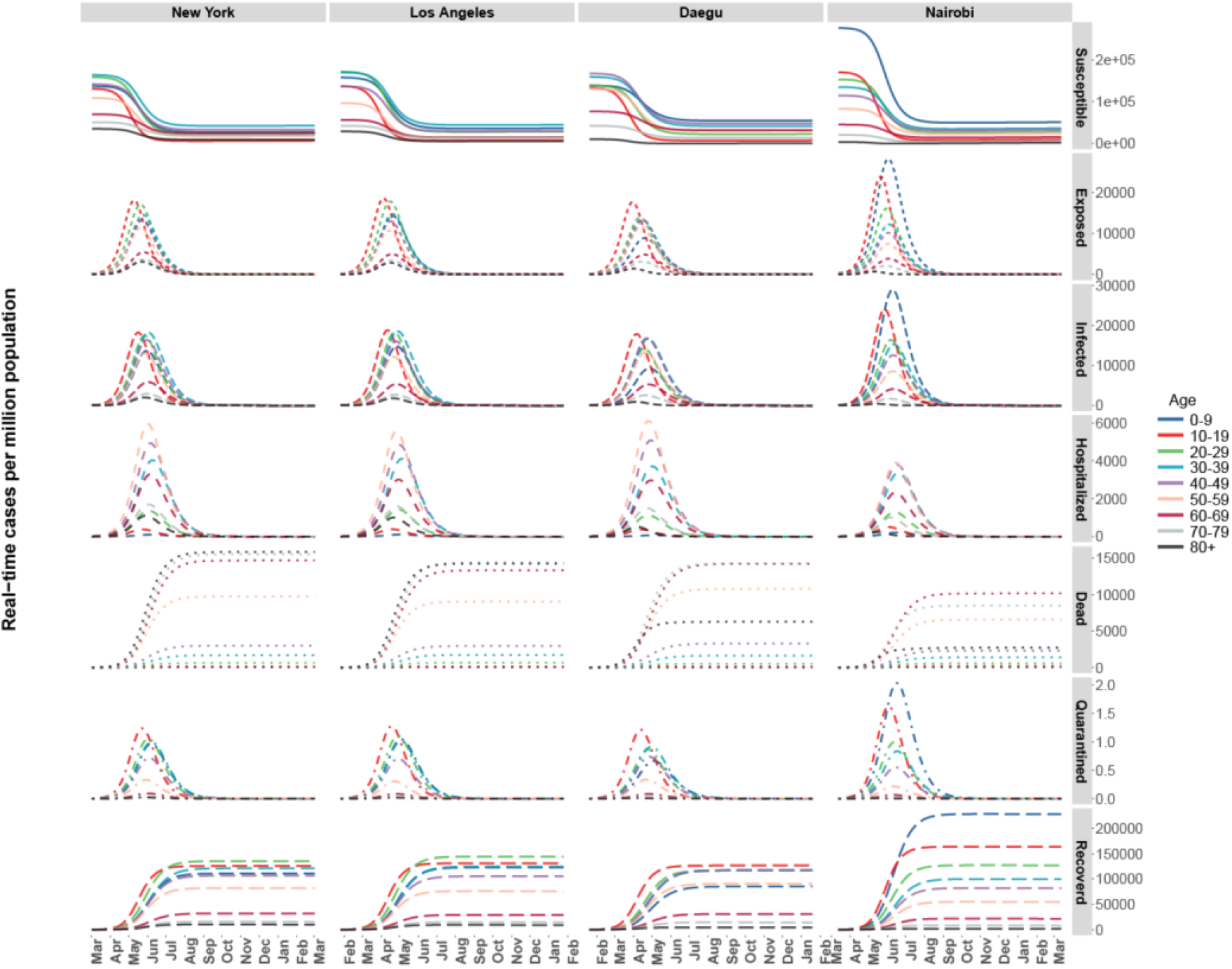
Real-time cases of different states under the risk of COVID-19 without policy interventions.

We find that school closure avoids 12%, 21%, 14% and 15% of infection in Nairobi, Daegu, New York and Los Angeles (Figure 4). A strategy of 50% of the population working from home only avoided 5% infection in Nairobi and 9%-10% in the other three cities. This clarifies how important it is to combine working remotely with other mitigation strategies to flatten the curve. An 80% population working from home strategy results (Scenario C) in 77% more avoided infection than that in Scenario B. A 50% reduction in other mobility helped Nairobi flatten the curve more effectively than the same degree of the working from home strategy. In contrast, a 10% daily quarantine rate averted (Scenario F) 45%-53% of all infections, which was more effective than a strict single social distancing strategy. The sustained combination of all four interventions together reduced the fatality rates by 96%-99% (Scenario G).

**Figure 4.**
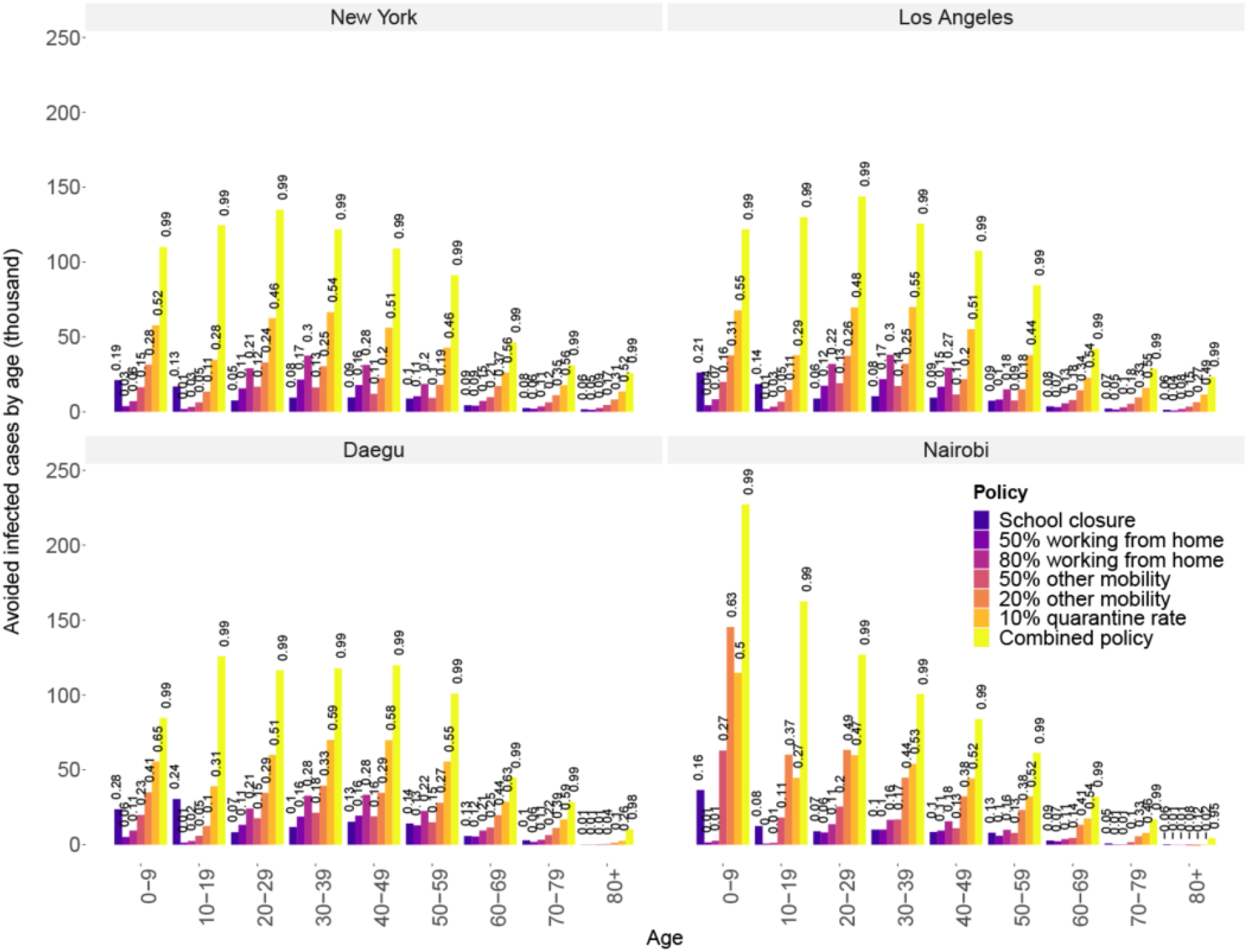
Avoided infected cases of the five policy interventions (Scenarios A – G in Table 2) by age in the four cities.

The effects of these five interventions also generated diverse effects for susceptible individuals in different ages. School closures resulted in largest impacts on the younger population aged from 0-19 and 50-59 years old population in four cities. The impact on the 50-59 year old group is an interesting interaction of the lower population exposure and the increasing susceptibility of this age group. A 50% work from home policy can avoid 14% of the infections in for the middle-age population (20-59 years old) in Daegu, Los Angeles and New York, while only avoid 9% infection for middle-age population in Nairobi since the transmission rates of working people were lower than the other three cities. A 50% reduction in other mobility can result in most avoided infection in older-age population (60 years and older) which accounted for 12%, 17%, 19% and 21% of the no intervention scenario.

**Table 2.**
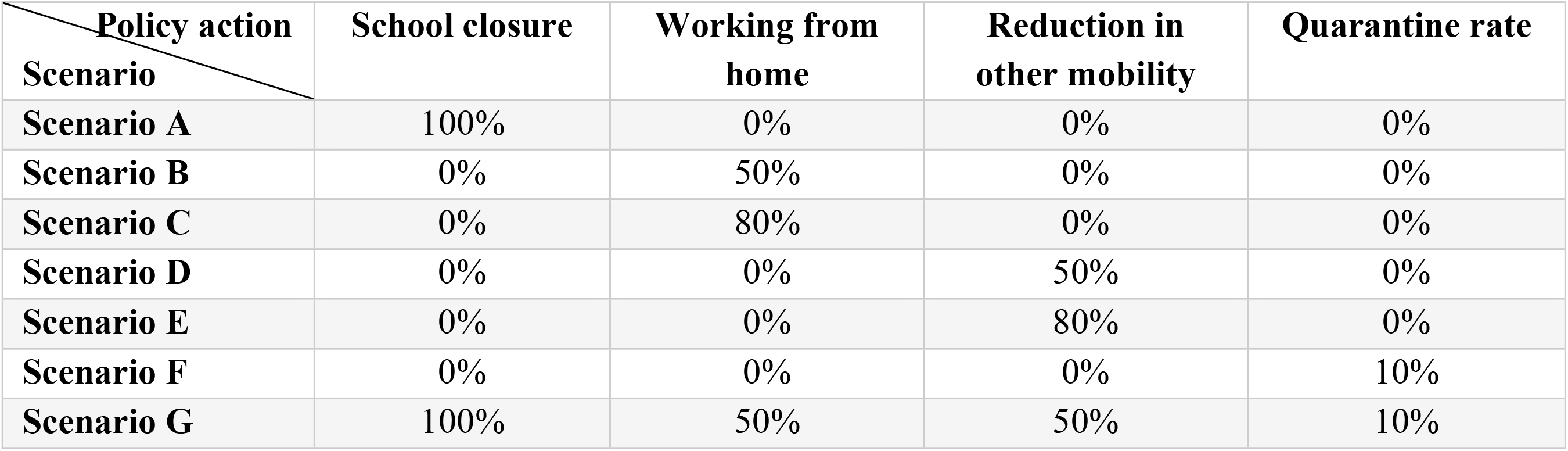
Scenario design for age-specific effects analysis.

The effectiveness of each intervention varied by the age of population. A 10% quarantine of the infected across all age groups was the most effective strategy in each of the four cities, which avoided 40%-50% of total infections for people who were 20-69 years old compared to the no intervention baseline. The quarantine strategy curbed the epidemic most quickly, however the spread of COVID-19 can then bounce back if we relaxed these interventions after 8 months (Figure 2a in Appendix 1).

A key finding of this work is that a combination of these four strategies could avoid 98%-99% over an 8 months intervention. It is important to take note of this finding, because as of this writing, most U. S. cities have been under quarantine for less than one month, and already political and social backlash against the strategies is growing in some communities and among some politicians. Under Scenario F and Scenario G, the four cities could be hit by second wave. Therefore, it is critical to maintain the interventions to avoid a second wave of COVID-19, which has now been mildly observed in China. We observe significant variations in the impacts of the different social distancing strategies, while the quarantine strategies (Scenario F and G) show are more uniform effect. With combination of these four interventions, limited age-specific effects were observed in the simulation. A moderate social distancing combined with high quarantine intervention (Scenario G) was effective enough to curb the spread of epidemic in 3 months (Appendix 1).

In the simulation of five interventions, a 10% quarantine rate avoided over 50% of the spread of the epidemic without totally shutting down the economy. Therefore, it is going to be beneficial for cities that are capable to diagnose exposed individuals and implement a detail epidemiological survey and then quarantine them strictly to avoid secondary transmission. However, social distancing should be combined to avoid fast hitting-back effect shown in Figure 5. This strategy together with moderate social distancing can effectively avoid strict social distancing and city lockdown interventions which could be costly to the economy.

**Figure 5.**
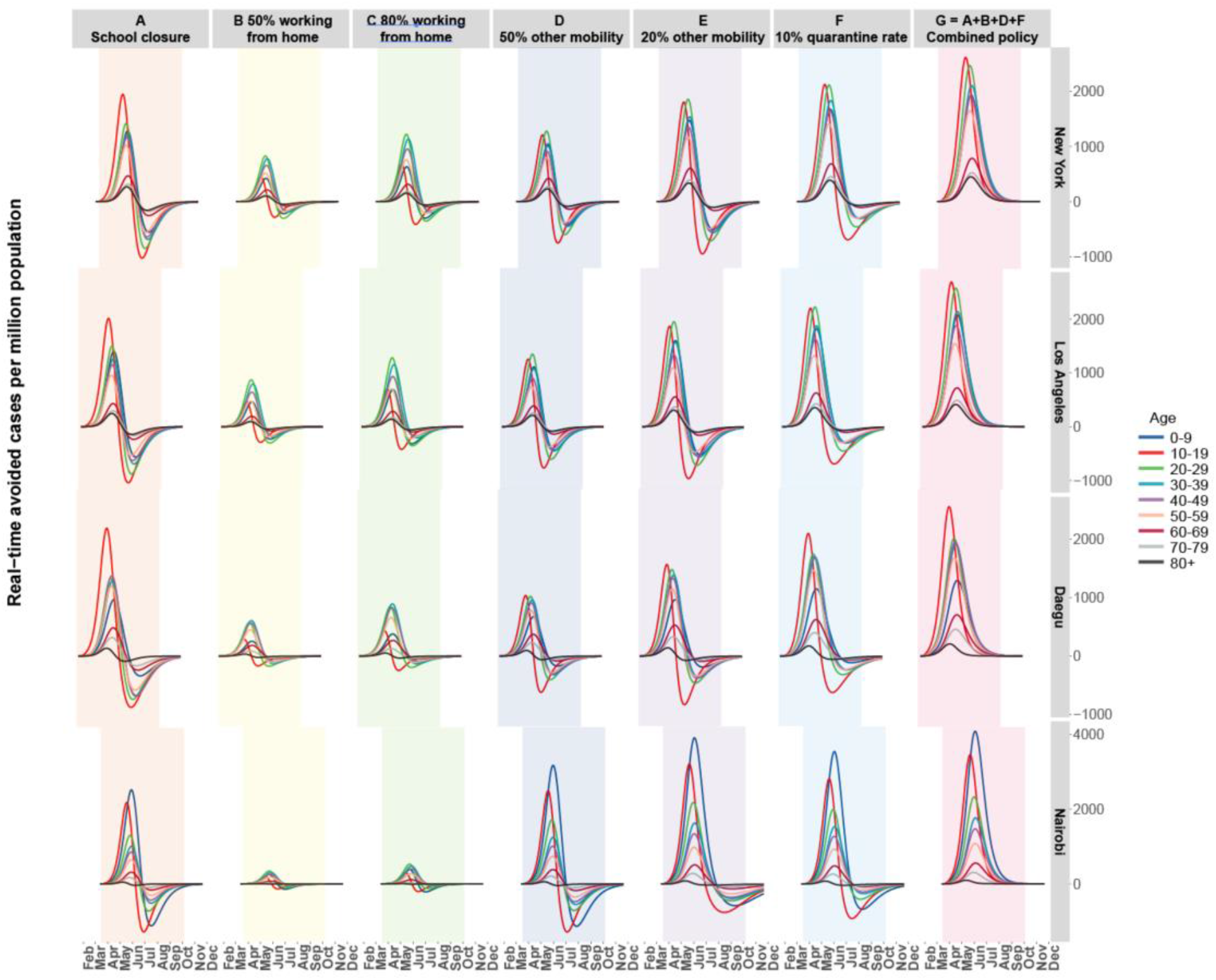
Time-series of the avoided cases distribution in the four cities by age. The shadowed regions represent the periods of interventions. Here the interventions take effects for 8 months since the intervention started after a week of the first case detected in each city.

**Figure 6.**
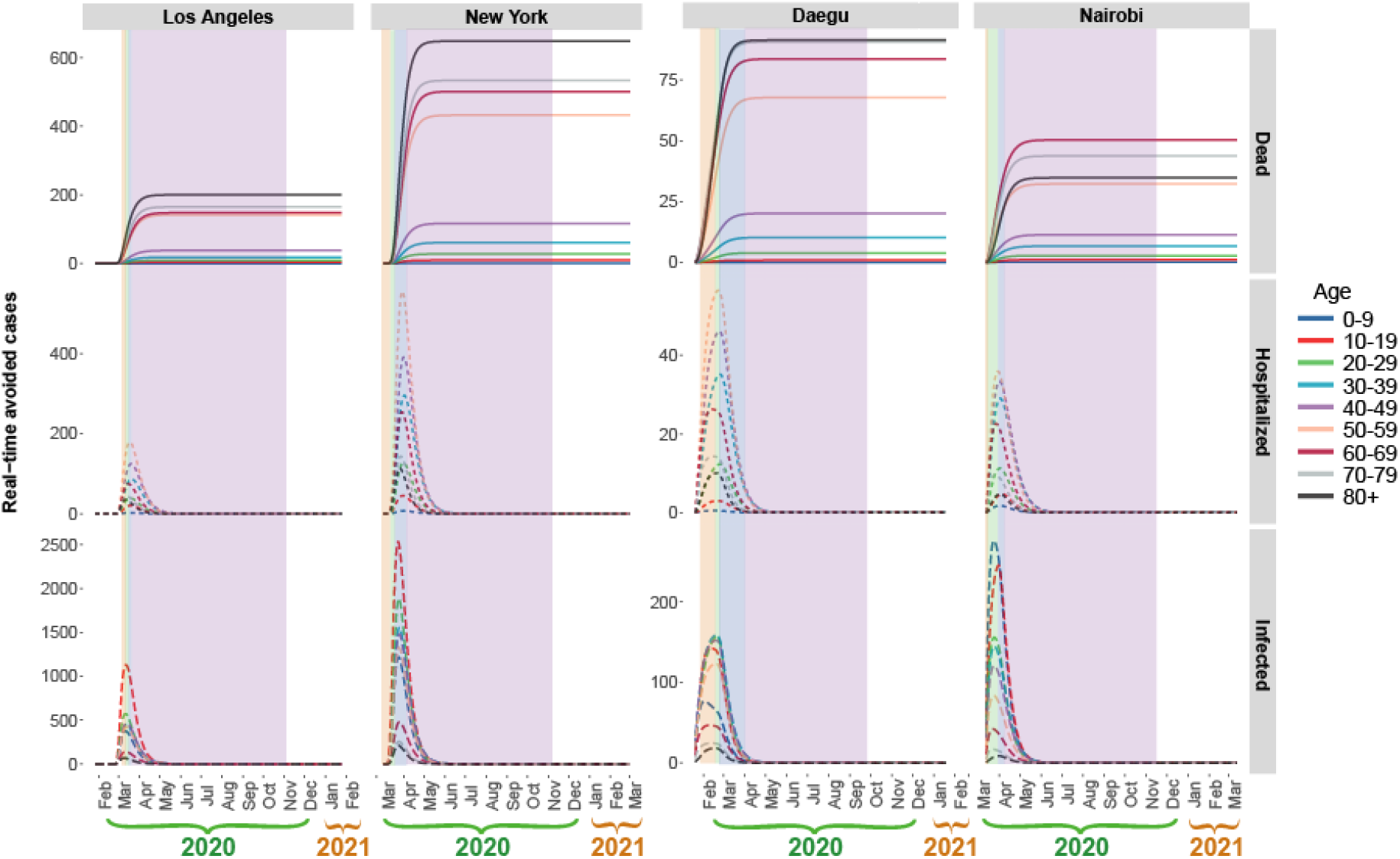
Age-specific effects of 1-week earlier intervention in four cities. The four colors of shadows represent four interventions and the widths of the shadows represent the duration of each intervention.

### 2. Age-specific benefits of intervention timing

Figure 6 shows the avoided dead, hospitalized and infected cases of COVID-19 under 1-week earlier interventions compared with the timing of their current policy interventions that were already implemented in the four cities.

An earlier intervention avoided 42%, 33%, 5%, and 16% deaths related to COVID-19 in Los Angeles, New York, Daegu and Nairobi. The earlier intervention benefited population aged 80 years and older the most in Nairobi, and Daegu, while the intervention avoided more deaths among population aged 10-19 years old in New York and Los Angeles. A similar trend can also be found in the containment effect of infection of the epidemic. By intervening 1-week earlier, Daegu avoids 250 cases of infection, which accounted for 5% of the infection in the business-as-usual (BAU) scenario (policy actions implemented in the real case). We find that Los Angeles and New York could have avoided 43% and 38% of infection if the interventions were introduced 1-week earlier. The earlier intervention strategy also mitigated the overwhelming requirement of hospital beds. Without implementing strict containment strategies, moving intervention earlier could help flatten the curve to certain degree depending on the timing of BAU interventions taking effects.

## Discussion

We developed, documented, and provided an open source model (available at https://github.com/HaoYinV/SEIR-HQD and at http://rael.berkeley.edu) which we utilized to examine the effects of different intervention strategies as nations manage risks to different age groups, and as sub-national regions work to curb the spread of COVID-19, and avoid the risks of a second reinfection wave. We tracked and modeled interventions in Daegu, South Korea, Nairobi, Kenya, and in Los Angeles and New York in the USA, to capture the effectiveness of these interactions between different public health interventions and population demographics. We explored the age-specific effects of COVID-19 interventions and their combination could differ depending on the age structure of susceptible population. We found that social distancing strategy in different location might generate stronger age-differential effects compared with the quarantine strategy.

Nairobi might have more infected cases than New York, Los Angeles and Daegu since younger people are relatively more active with higher transmission rates in no intervention scenario, however, there were more cases of fatality and hospitalization in the three cities with an ageing population. Under five intervention scenarios, we found that school closure avoided more infection of population aged 0-19 years old, while working from home and reduction in other mobility had more averted infection in middle aged population. This leads to significantly reductions in death rates, which a strongly skewed to the younger and middle population, more actions are needed for the older population.

In addition, we find the need to guard by sustaining interventions in Nairobi that could otherwise experience a second wave of infections that would hit faster than in the other three cities owing to the higher transmission rates in younger people. Cities with older population should take actions by reducing mobility in the older population and encouraging younger population to go to work combining with social distancing strategy and growing quarantine rates. We showed that it was possible to contain the pandemic with strategy of high quarantine rate together with moderate social distancing in working and other mobility which could largely avoid the costly influence on the national and global economy. Earlier intervention could help flatten the curve in Los Angeles and New York the most due to the relative slow actions since the first case was detected than other two cities. We did not find the substantial difference of the early intervention on population in different ages in the simulations.

There is limited study showing that susceptibility and transmissibility vary with the age of the population at risk.^9^ The SEIR-HQD model was based on assumption that the transmissibility only depends on the contact rates and population sizes in each age group. In our model, we assume the quarantine only happened to people who showed symptoms, whereas, the quarantine strategy could even expand to the exposed but have no symptom yet. Therefore, an expanding quarantine intervention might have even larger effects than the results from our simulations. Since there was no city-specific fatality rates in each city, we adopted the age-specific fatality of China in four cities that was adjusted for censoring demographic information without the consideration of the medical capacity and facility.^1^ In addition, other environmental risks, such as air pollution, might have significant impacts on the fatality rate of COVID-19 geographically.^22^ A unified age-specific fatality rate might over- or underestimate the estimates of deaths in the four cities. In our model, we did not consider the population dynamics during the pandemic. Next step, we will further incorporate the impacts of population movement and air quality inside the SEIR-HQD model.

In conclusion, the combination of moderate social distancing and high quarantine rate could avoid over 90% of the infection and flatten the epidemic curve in a short period of time. Social distancing might have age-specific effects; therefore, cities with a high proportion of ageing population should reduce their mobility and protect them from younger population who tend to have higher transmission rates. Cities with younger population should be more careful about the second hit, while cities with ageing population should prepare more hospital beds and resources.

## Data Availability

The modeling code and estimation process are available online at https://github.com/HaoYinV/SEIR-HQD and http://rael.berkeley.edu

## Open Access Code and Data sharing

The data used in this study is publicly available.^12^ The modeling code and estimation process are available online at https://github.com/HaoYinV/SEIR-HQD.

## Acknowledgements

HY was funded by the National Natural Science Foundation of China (grant 71904104) and the China Postdoctoral Science Foundation (grant 2019M650726). ZL acknowledges funding from Qiushi Foundation, the Resnick Sustainability Institute at California Institute of Technology and the National Natural Science Foundation of China (grant 71874097 and 41921005). DMK gratefully acknowledges the support of the Zaffaroni Family Foundation, the Karsten Family Foundation, and US NSF CyberSEES Grant #1539585 and NSF InFEWS grant DGE #1633740.

## Appendix 1

### 1 Population age structure

Compared with New York, Los Angeles and Daegu, the population in Nairobi was much younger (Figure 1a).

**Figure 1a.**
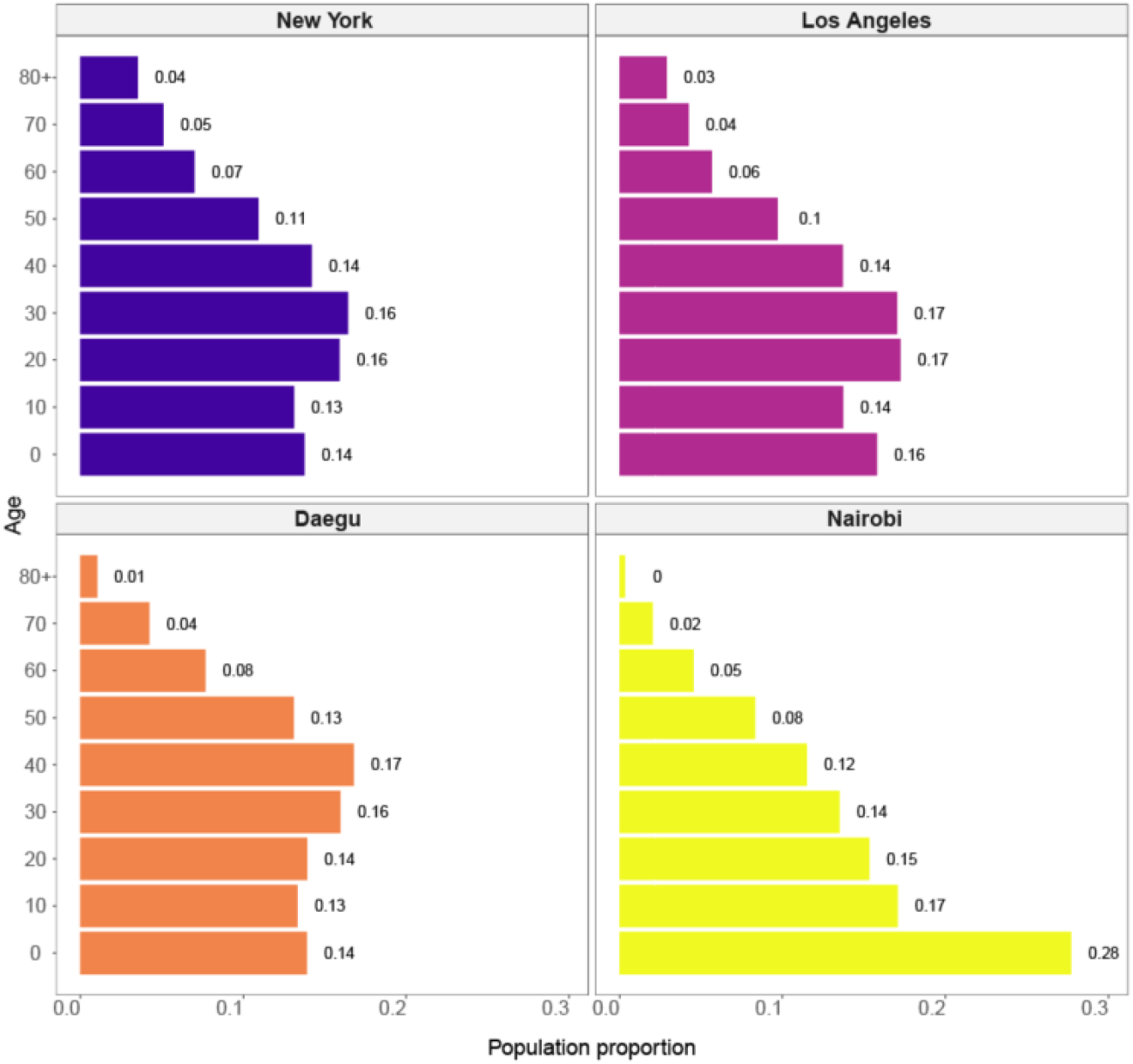
Population age structure in four cities

### 2 Birth and death rates

**Table 1a.** Birth rates in the four cities

| City | New York | Los Angeles | Daegu | Nairobi |
| --- | --- | --- | --- | --- |
| Birth rate per 1000 people/year | 11.8 | 11.8 | 7.4 | 29.3 |

In our simulation we also consider the dynamics of population birth and death rates without the risks of COVID-19. In addition, the population will transit from one age group to another over time of the simulation.

**Table 2a.** Death rates by age in the four cities

| City | New York | Los Angeles | Daegu | Nairobi |
| --- | --- | --- | --- | --- |
| Age |  |  |  |  |
| 0-9 | 0.0013 | 0.0013 | 0.0003 | 0.0036 |
| 10-19 | 0.0004 | 0.0004 | 0.0003 | 0.0017 |
| 20-29 | 0.0010 | 0.0010 | 0.0005 | 0.0036 |
| 30-39 | 0.0016 | 0.0016 | 0.0010 | 0.0064 |
| 40-49 | 0.0030 | 0.0030 | 0.0023 | 0.0103 |
| 50-59 | 0.0064 | 0.0064 | 0.0050 | 0.0185 |
| 60-69 | 0.0126 | 0.0126 | 0.0133 | 0.0407 |
| 70-79 | 0.0266 | 0.0266 | 0.0460 | 0.0930 |
| 80+ | 0.0709 | 0.0709 | 0.0853 | 0.1346 |

Table 3a Hospitalization rate by age in the four cities

Table 4a Fatality rate by age in the four cities

### 3 Transmission rates

**Table 5a.**
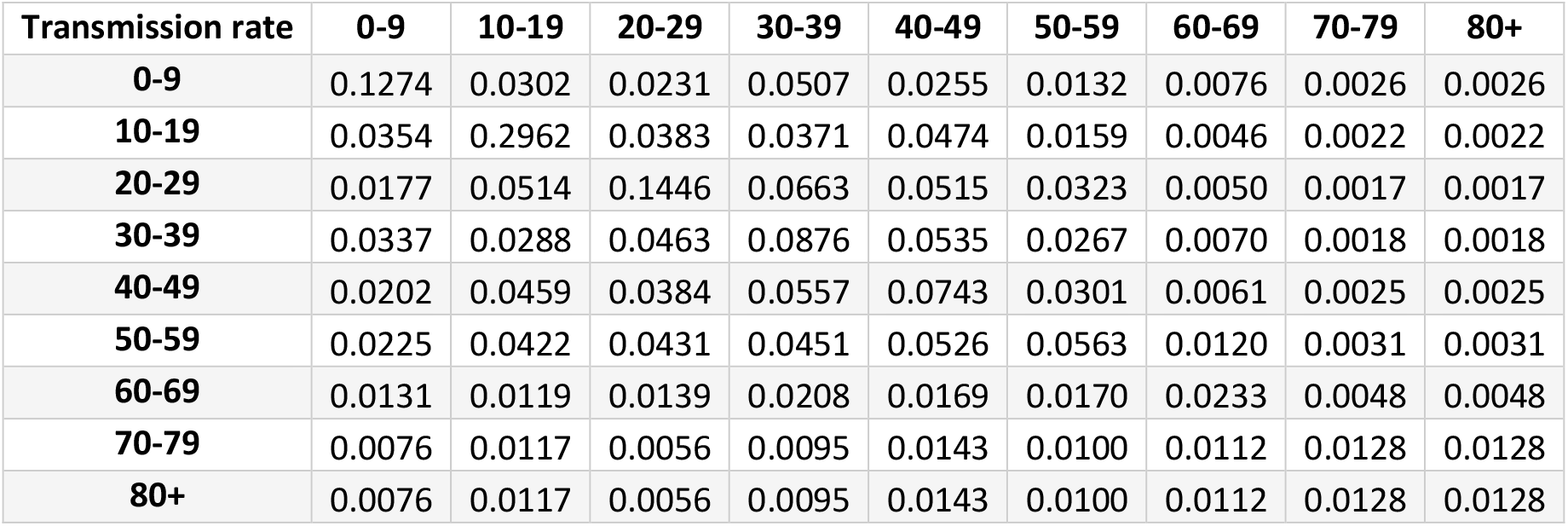
Transmission rates by age in New York

| Transmission rate | 0-9 | 10-19 | 20-29 | 30-39 | 40-49 | 50-59 | 60-69 | 70-79 | 80+ |
| --- | --- | --- | --- | --- | --- | --- | --- | --- | --- |
| 0-9 | 0.1274 | 0.0302 | 0.0231 | 0.0507 | 0.0255 | 0.0132 | 0.0076 | 0.0026 | 0.0026 |
| 10-19 | 0.0354 | 0.2962 | 0.0383 | 0.0371 | 0.0474 | 0.0159 | 0.0046 | 0.0022 | 0.0022 |
| 20-29 | 0.0177 | 0.0514 | 0.1446 | 0.0663 | 0.0515 | 0.0323 | 0.0050 | 0.0017 | 0.0017 |
| 30-39 | 0.0337 | 0.0288 | 0.0463 | 0.0876 | 0.0535 | 0.0267 | 0.0070 | 0.0018 | 0.0018 |
| 40-49 | 0.0202 | 0.0459 | 0.0384 | 0.0557 | 0.0743 | 0.0301 | 0.0061 | 0.0025 | 0.0025 |
| 50-59 | 0.0225 | 0.0422 | 0.0431 | 0.0451 | 0.0526 | 0.0563 | 0.0120 | 0.0031 | 0.0031 |
| 60-69 | 0.0131 | 0.0119 | 0.0139 | 0.0208 | 0.0169 | 0.0170 | 0.0233 | 0.0048 | 0.0048 |
| 70-79 | 0.0076 | 0.0117 | 0.0056 | 0.0095 | 0.0143 | 0.0100 | 0.0112 | 0.0128 | 0.0128 |
| 80+ | 0.0076 | 0.0117 | 0.0056 | 0.0095 | 0.0143 | 0.0100 | 0.0112 | 0.0128 | 0.0128 |

**Table 6a.**
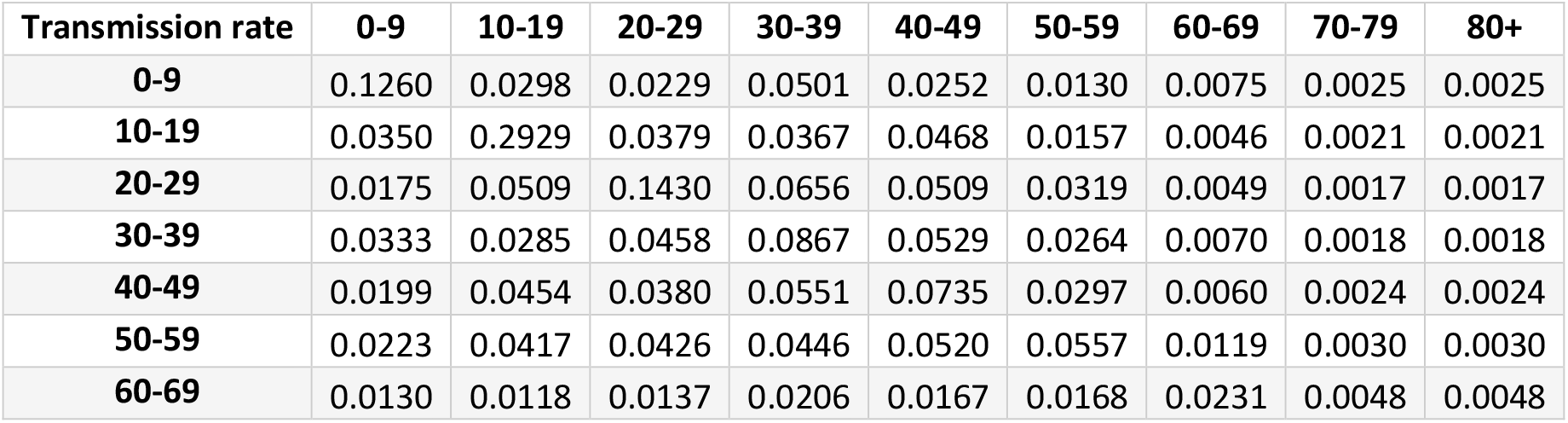
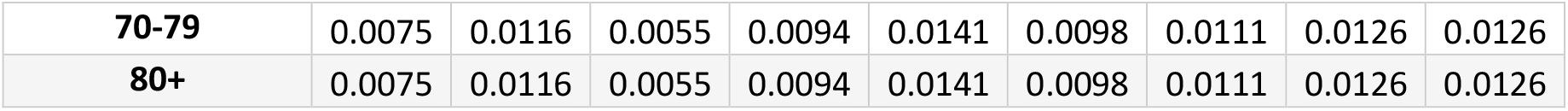
Transmission rates by age in Los Angeles

| Transmission rate | 0-9 | 10-19 | 20-29 | 30-39 | 40-49 | 50-59 | 60-69 | 70-79 | 80+ |
| --- | --- | --- | --- | --- | --- | --- | --- | --- | --- |
| 0-9 | 0.1260 | 0.0298 | 0.0229 | 0.0501 | 0.0252 | 0.0130 | 0.0075 | 0.0025 | 0.0025 |
| 10-19 | 0.0350 | 0.2929 | 0.0379 | 0.0367 | 0.0468 | 0.0157 | 0.0046 | 0.0021 | 0.0021 |
| 20-29 | 0.0175 | 0.0509 | 0.1430 | 0.0656 | 0.0509 | 0.0319 | 0.0049 | 0.0017 | 0.0017 |
| 30-39 | 0.0333 | 0.0285 | 0.0458 | 0.0867 | 0.0529 | 0.0264 | 0.0070 | 0.0018 | 0.0018 |
| 40-49 | 0.0199 | 0.0454 | 0.0380 | 0.0551 | 0.0735 | 0.0297 | 0.0060 | 0.0024 | 0.0024 |
| 50-59 | 0.0223 | 0.0417 | 0.0426 | 0.0446 | 0.0520 | 0.0557 | 0.0119 | 0.0030 | 0.0030 |
| 60-69 | 0.0130 | 0.0118 | 0.0137 | 0.0206 | 0.0167 | 0.0168 | 0.0231 | 0.0048 | 0.0048 |
| **70-79** | 0.0075 | 0.0116 | 0.0055 | 0.0094 | 0.0141 | 0.0098 | 0.0111 | 0.0126 | 0.0126 |
| **80+** | 0.0075 | 0.0116 | 0.0055 | 0.0094 | 0.0141 | 0.0098 | 0.0111 | 0.0126 | 0.0126 |

**Table 7a.** Transmission rates by age in Daegu

| **Transmission rate** | **0-9** | **10-19** | **20-29** | **30-39** | **40-49** | **50-59** | **60-69** | **70-79** | **80+** |
| --- | --- | --- | --- | --- | --- | --- | --- | --- | --- |
| **0-9** | 0.0811 | 0.0186 | 0.0138 | 0.0343 | 0.0218 | 0.0096 | 0.0048 | 0.0020 | 0.0020 |
| **10-19** | 0.0284 | 0.3078 | 0.0308 | 0.0308 | 0.0470 | 0.0168 | 0.0037 | 0.0023 | 0.0023 |
| **20-29** | 0.0123 | 0.0417 | 0.1265 | 0.0615 | 0.0500 | 0.0351 | 0.0059 | 0.0020 | 0.0020 |
| **30-39** | 0.0322 | 0.0254 | 0.0437 | 0.0933 | 0.0554 | 0.0283 | 0.0089 | 0.0025 | 0.0025 |
| **40-49** | 0.0199 | 0.0463 | 0.0366 | 0.0577 | 0.0808 | 0.0317 | 0.0067 | 0.0032 | 0.0032 |
| **50-59** | 0.0190 | 0.0414 | 0.0423 | 0.0460 | 0.0551 | 0.0575 | 0.0113 | 0.0038 | 0.0038 |
| **60-69** | 0.0096 | 0.0090 | 0.0125 | 0.0225 | 0.0176 | 0.0163 | 0.0189 | 0.0044 | 0.0044 |
| **70-79** | 0.0069 | 0.0108 | 0.0056 | 0.0114 | 0.0154 | 0.0111 | 0.0106 | 0.0122 | 0.0122 |
| **80+** | 0.0069 | 0.0108 | 0.0056 | 0.0114 | 0.0154 | 0.0111 | 0.0106 | 0.0122 | 0.0122 |

**Table 8a.** Transmission rates by age in Nairobi

| **Transmission rate** | **0-9** | **10-19** | **20-29** | **30-39** | **40-49** | **50-59** | **60-69** | **70-79** | **80+** |
| --- | --- | --- | --- | --- | --- | --- | --- | --- | --- |
| **0-9** | 0.2020 | 0.0541 | 0.0351 | 0.0397 | 0.0160 | 0.0075 | 0.0040 | 0.0012 | 0.0012 |
| **10-19** | 0.0642 | 0.2993 | 0.0419 | 0.0278 | 0.0223 | 0.0069 | 0.0025 | 0.0010 | 0.0010 |
| **20-29** | 0.0296 | 0.0559 | 0.1129 | 0.0483 | 0.0286 | 0.0131 | 0.0034 | 0.0005 | 0.0005 |
| **30-39** | 0.0378 | 0.0277 | 0.0358 | 0.0491 | 0.0317 | 0.0119 | 0.0031 | 0.0004 | 0.0004 |
| **40-49** | 0.0247 | 0.0341 | 0.0240 | 0.0338 | 0.0334 | 0.0132 | 0.0029 | 0.0004 | 0.0004 |
| **50-59** | 0.0197 | 0.0249 | 0.0188 | 0.0208 | 0.0230 | 0.0129 | 0.0036 | 0.0006 | 0.0006 |
| **60-69** | 0.0129 | 0.0111 | 0.0075 | 0.0099 | 0.0080 | 0.0062 | 0.0025 | 0.0007 | 0.0007 |
| **70-79** | 0.0076 | 0.0120 | 0.0024 | 0.0031 | 0.0036 | 0.0031 | 0.0021 | 0.0012 | 0.0012 |
| **80+** | 0.0076 | 0.0120 | 0.0024 | 0.0031 | 0.0036 | 0.0031 | 0.0021 | 0.0012 | 0.0012 |

### 4 Real-time infected cases in 7 scenarios

**Figure 2a.**
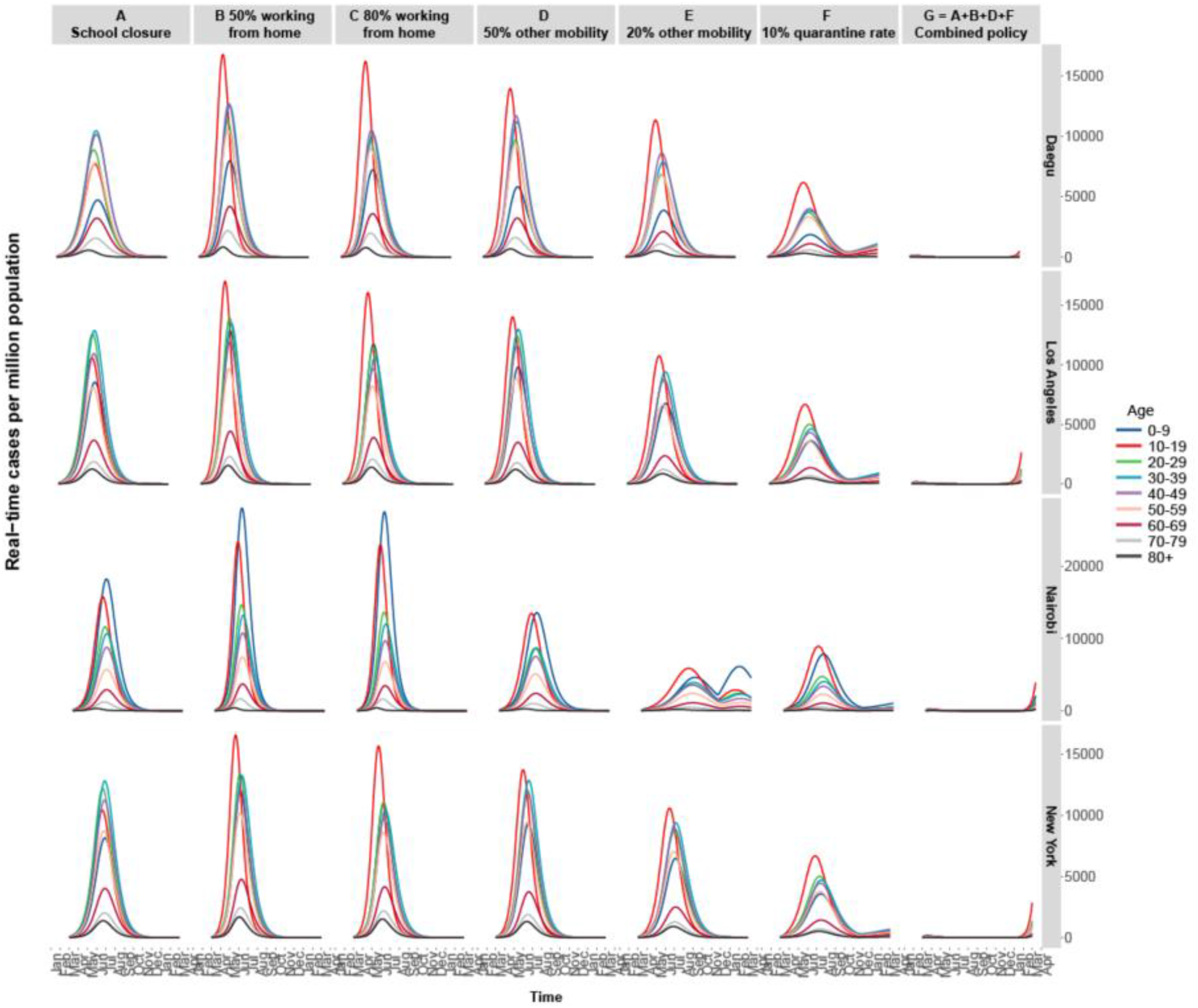
The infected cases of four cities in 7 scenarios

### 5 Policy database in each city

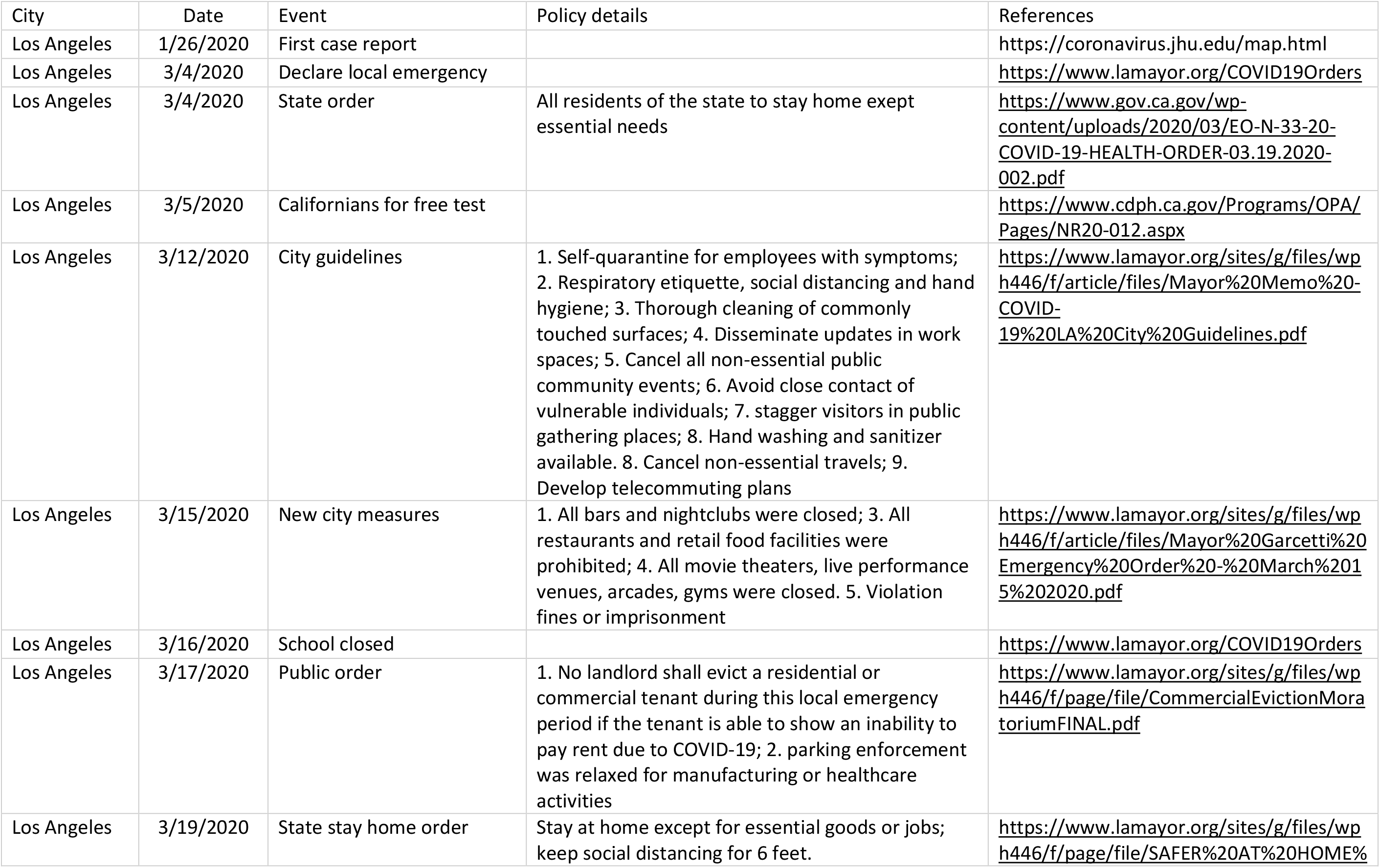
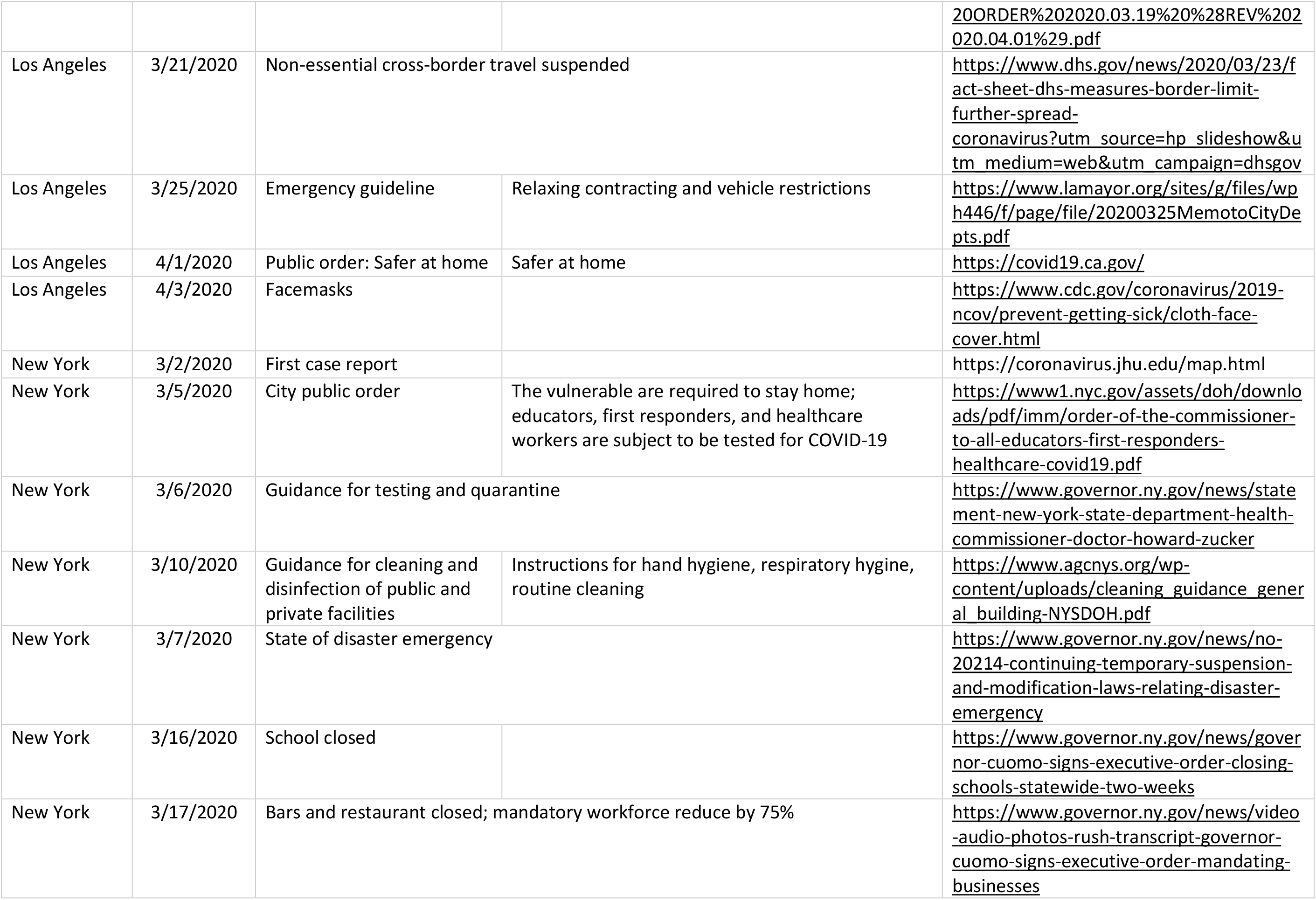
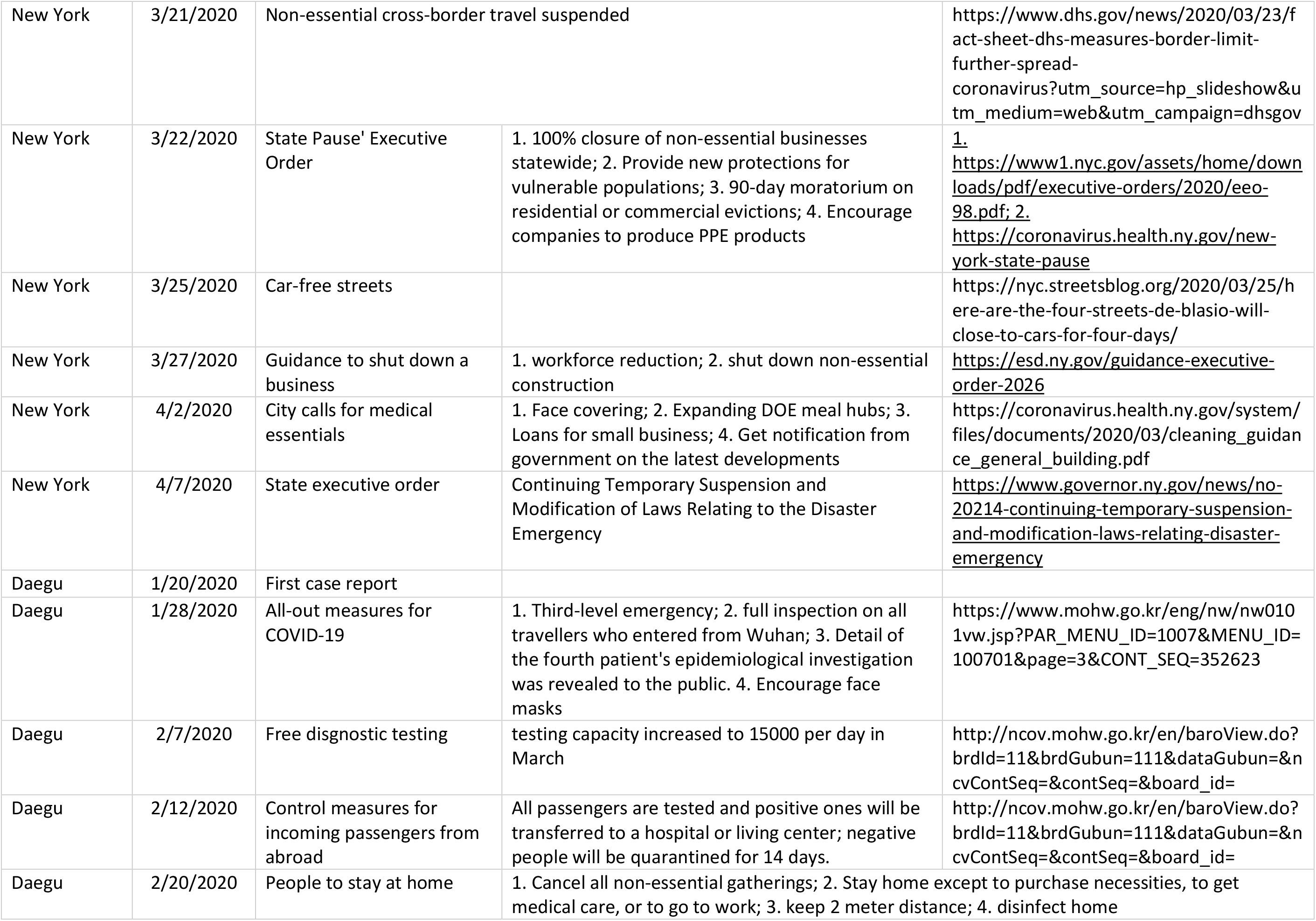
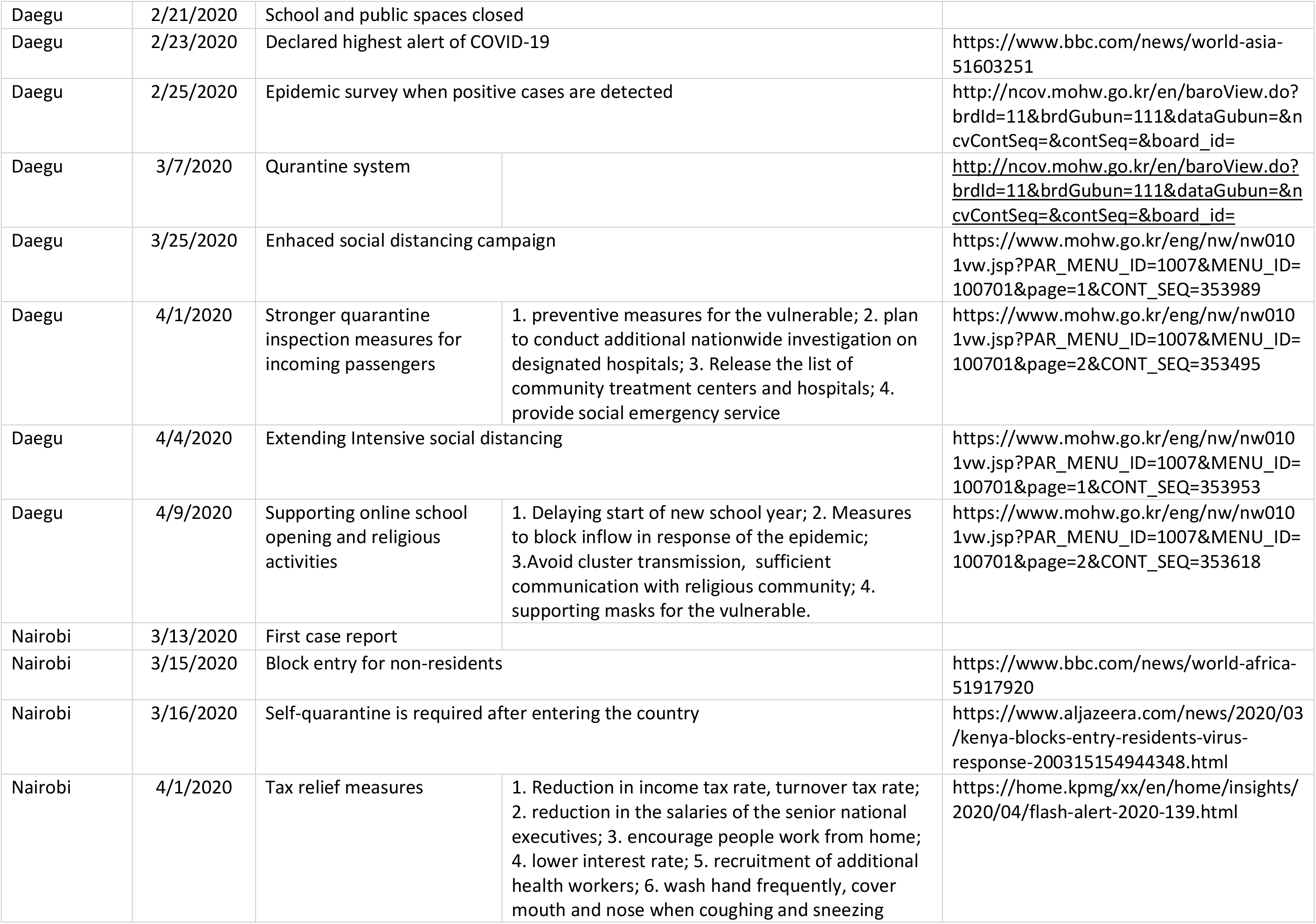
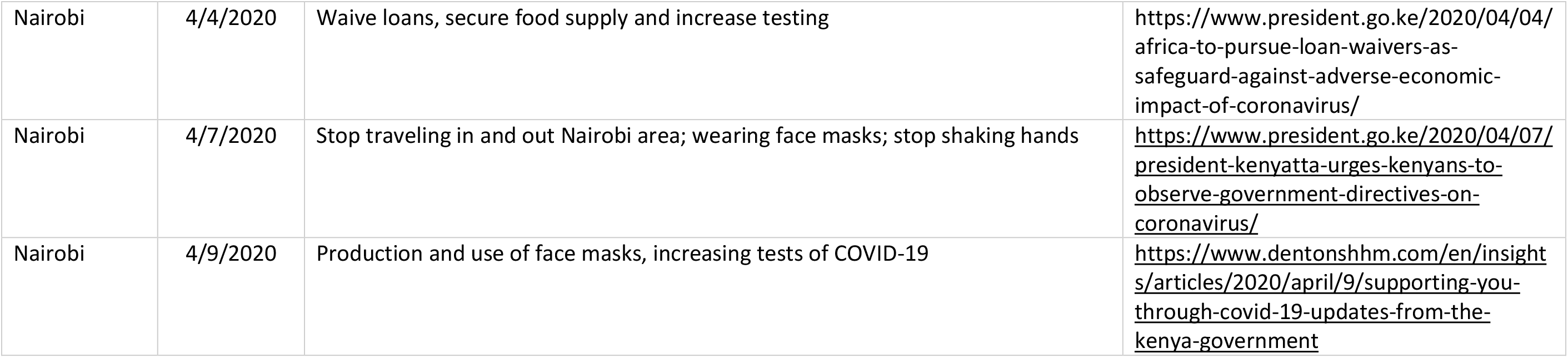

